# Substance Use is Not Associated with Antidepressant Response to Transcranial Magnetic Stimulation

**DOI:** 10.64898/2026.09.01.26361949

**Authors:** Jordyn Chesley, Kathryn Biernacki, Jordan Vanleuven, John Doran, Idil Yazgan, Gulcan Yildiz, Daniel Alejandro Gonzalez, Shannon Young Wagner, Kaylee LeBaron, Emily Marrero, Tagalsir Osama, Simon Vandekar, Heather Burrell Ward

## Abstract

**Background:** Substance use is common among individuals with depression. Transcranial magnetic stimulation (TMS) is an effective treatment for depression, but current clinical guidelines have discouraged TMS treatment for individuals with depression and co-occurring substance use given concerns for limited efficacy. However, limited data exists on whether substance use affects response to TMS.

**Methods:** Using electronic health record data from patients who received a standard course of TMS for major depressive disorder at an academic medical center, we investigated associations between substance use frequency and response to TMS, defined as change in Patient Health Questionnaire-9 (PHQ-9) scores. Substance use frequency was extracted for alcohol, cannabis, nicotine, stimulants, benzodiazepines, opioids, inhalants, psychedelics, and other drugs. We performed ANCOVA and multiple regression analyses to predict change in PHQ-9 score based on substance use frequency, controlling for pre-TMS PHQ-9 score, age, sex, and number of TMS sessions received.

**Results:** We extracted data from 219 TMS courses. Alcohol was the substance used most commonly (34.2%), followed by prescription benzodiazepines (28.3%), and prescription stimulants (21.0%). Across all substance categories, substance use was not associated with change in PHQ-9 score (all *p’s* > 0.05, Cohen’s d=0.00 to 0.30). In multiple regression models to compare individual levels of substance use frequency (e.g., daily use vs. no use), level of substance use was not associated with change in PHQ-9 score (all *p’s* > 0.05). The range of plausible effects of substance use frequency on PHQ-9 change was generally below the minimal clinically important difference for PHQ-9, suggesting substance use was unlikely to have a meaningful clinical effect on antidepressant response to TMS.

**Conclusions:** Low to moderate substance use does not have a clinically significant effect on antidepressant response to TMS. Low-level substance use should not exclude individuals with depression from receiving TMS.

## Introduction

Depressive disorders are among the leading causes of disability worldwide (GBD Mental Disorders Collaborators, 2022), with major depressive disorder (MDD) affecting more than 185 million people globally (Yan et al., 2024). Although several effective treatments for depression are available, many individuals do not achieve adequate symptom improvement (Dodd et al., 2021). At the same time, substance use is common among people with MDD (Hunt, Malhi, Lai, & Cleary, 2020; Volkow, 2004), and the use of both licit and illicit substances may impact the effectiveness of depression treatments (Howland et al., 2009; Nunes & Levin, 2004). However, the extent to which substance use affects treatment efficacy remains poorly understood. A better understanding of the relationship between substance use and treatment response is needed to determine how substance use affects treatment efficacy and to identify opportunities for optimizing treatment approaches for individuals with MDD.

Transcranial magnetic stimulation (TMS) is an FDA-approved and now commonly utilized treatment for MDD (Cohen, Bikson, Badran, & George, 2022). TMS delivers brief electromagnetic pulses to targeted cortical regions, most commonly the dorsolateral prefrontal cortex (dlPFC), to modulate cortical excitability. This stimulation is thought to enhance blunted cortical activity and promote neurotransmitter release, thereby improving depressive symptoms (Avissar et al., 2017; Baeken & De Raedt, 2011; Pogarell et al., 2006). Several TMS protocols have been approved for the treatment of depression, including high-frequency and low-frequency TMS, as well as intermittent theta burst stimulation (iTBS). These protocols can be administered as conventional once-daily sessions or as accelerated treatment schedules in which multiple stimulation sessions are delivered within a single day (Chen et al., 2025). Extensive evidence supports the safety of these approaches in individuals with MDD (George, 2010). However, the efficacy of TMS is variable, and many patients do not achieve a meaningful clinical benefit. Current estimates suggest that approximately 50% of patients experience a clinically significant response to TMS, while only about 30% achieve remission (Chen et al., 2025). Consequently, a substantial treatment gap remains, which may be influenced by a range of factors, including concurrent substance use.

Concurrent substance use is common for people diagnosed with MDD (Hunt et al., 2020) and may represent an important, but understudied, factor influencing response to TMS. Even without a diagnosis of substance use disorder, people with depression will use substances at a higher rate than the general population (Substance Abuse and Mental Health Services Administration, 2021). Drugs of abuse impact cortical excitability (Barr, Fitzgerald, Farzan, George, & Daskalakis, 2008; Leyrer-Jackson, Hood, & Olive, 2021), which in turn may impact the ability of TMS to modulate brain activity. Most commonly, alcohol and benzodiazepine use have well-known effects on cortical excitability (Koob & Volkow, 2016). For example, cannabis use disorder and methamphetamine use have both been associated with increased cortical excitability (Khedr, Elserogy, & Ahmed, 2025; Khedr, Elserogy, Goda, & Fawzy, 2026). Correspondingly, young people with cannabis use disorder have been shown to have a blunted cortical response to TMS (Martin Rodriguez et al., 2021). In contrast, heroin use has been linked to impaired cortical plasticity in response to TMS (Shen, Cao, Shan, Dai, & Yuan, 2017), a mechanism thought to be important for the therapeutic effects of TMS (Jannati, Oberman, Rotenberg, & Pascual-Leone, 2023). There are also concerns that alterations in cortical excitability can predispose to seizure, a rare but known potential side effect of TMS. However, despite evidence that substance dependence can alter cortical excitability and plasticity, the impact of substance use that does not meet criteria for a substance use disorder on the therapeutic effects of TMS for depression remains unknown.

Clarifying the relationship between substance use and TMS treatment outcomes is essential for developing evidence-based clinical recommendations and ensuring equitable access to care. Efforts to provide clinical guidance for the management of concurrent substance use during TMS treatment for psychiatric disorders have suggested that clinicians encourage complete abstinence before initiating treatment given concerns about the potential for decreased safety and efficacy (Tang et al., 2025). In practice, however, complete abstinence from substances may not be feasible or realistic for many patients (American Society of Addiction Medicine, 2024). At the same time, substantial gaps remain in the treatment of depression, with many individuals not receiving adequate care (Olfson, Blanco, & Marcus, 2016). One contributing factor may be the stigmatization of substance use within healthcare settings, which can reduce clinicians’ willingness to engage and treat individuals who use drugs or alcohol (Fong et al., 2021). Given the existing treatment gap for depression and the continued underutilization of TMS despite evidence supporting its effectiveness in real-world clinical settings (Bastiaens et al., 2024), it is important to clarify whether and how substance use influences treatment outcomes. Identifying predictors of TMS response will enable clinicians to make more informed treatment recommendations and may help expand access to care for patients who might otherwise be excluded from treatment.

To better characterize the relationship between substance use and the efficacy of TMS for the treatment of depression, this study examined how substance use impacted change in patient Health Questionnaire-9 (PHQ-9) scores, a routinely collected measure of depressive symptom severity, before to after a course of TMS treatment. To determine clinical significance, we used a change in PHQ-9 score of 5, which has been established as the minimal clinically important difference at the individual patient level (Löwe, Unützer, Callahan, Perkins, & Kroenke, 2004). We hypothesized that more frequent substance use would be associated with attenuated antidepressant response to TMS. By understanding the impact of substance use on TMS-related changes in depressive symptoms, this study aims to guide clinical decision-making and optimize clinical outcomes with TMS.

## Methods

### Study design and setting

This was a retrospective cohort study conducted at Vanderbilt University Medical Center, an academic medical center in Tennessee. The study utilized the electronic medical records (EMR) to extract data from patients who received TMS treatment for MDD between 7/11/2011 and 9/29/25. TMS was administered using a figure-of-eight coil with either a Neurostar or MagVenture stimulator. The study protocol was approved by the VUMC Institutional Review Board (IRB # 251256), with a waiver of informed consent granted given the retrospective nature of the study.

### Study population

Eligible patients were adults aged 18 years and older who (1) had a documented diagnosis of MDD (via diagnosis code or clinical diagnosis); (2) who had both PHQ-9 scores available pre- and post-TMS treatment, and (3) had documentation of substance use status. Exclusion criteria included (1) missing PHQ-9 data, (2) no diagnosis of MDD, or (3) missing documentation of substance use.

### Data collection

Data was extracted via Research Derivative, a data warehouse from the Vanderbilt Institute of Clinical and Translational Research (VICTR). A list of patients who had TMS Current Procedural Terminology (CPT) codes in their record was obtained via Research Derivative. TMS status was then determined via chart review. Through manual extraction, PHQ-9 scores were obtained before, during, and after a course of TMS.

Substance use status was extracted using a combination of ICD-10 diagnosis codes and clinical notes. After identification of participants, frequency of substance use prior to receiving TMS was extracted for the following substances: alcohol, cannabis, nicotine, stimulants (prescribed and illicit), benzodiazepines (prescribed and illicit), opioids (prescribed and illicit), inhalants, psychedelics, and other drugs. Frequency of use was categorized as: no use, less than weekly, more than weekly, and daily or more.

### Outcome measures

The primary outcome of the study was the change in PHQ-9 scores from pre-TMS to end of TMS treatment (post final TMS session).

### Statistical analyses

Descriptive statistics were used to characterize the study sample and patterns of substance use. We performed ANCOVAs for each substance category to determine the main effect of substance use frequency on change in PHQ-9 score (post-treatment – pre-treatment) in a model controlling for age, sex, number of TMS sessions, and pre-TMS PHQ-9 score. To compare individual levels of substance use frequency (i.e., daily use vs. no use), we used multivariable linear regression to evaluate the effect of substance use frequency on change in PHQ-9 score controlling for age, sex, number of TMS sessions, and pre-TMS PHQ-9 score for each substance type.

## Results

We identified a total of 219 total TMS courses with 212 unique patients. The sample had a mean age of 47.0 (SD 15.6), was 56.6% female with a mean baseline PHQ-9 score of 17.6 (SD 4.8), indicating moderate depression. Individuals received an average of 32.9 (SD 8.7) TMS sessions. Alcohol was the substance used most commonly (31.1%), followed by prescription benzodiazepines (27.8%), and prescription stimulants (21.0%). See Table 1 for patient characteristics. No individuals reported illicit stimulant use or inhalant use.

**Table 1.** Demographics.

| **Variable** | **MDD (n=219)** |
| --- | --- |
| Age, mean (SD) | 47.0 (15.6) |
| Female, n (% female) | 124 (56.6) |
| PHQ-9, mean (SD) | 17.6 (4.8) |
| Mean # of TMS sessions, (SD) | 32.9 (8.7) |
| **Race n, (%)** |  |
| Asian | 1 (0.5) |
| Black or African American | 8 (3.7) |
| White | 187 (85.4) |
| Unknown | 7 (3.2) |
| More than one Race | 7 (3.2) |
| No Answer | 9 (4.1) |
| **Ethnicity n, (%)** |  |
| Hispanic/Latino | 9 (4.1) |
| Not Hispanic or Latino | 194 (88.6) |
| Unknown | 16 (7.3) |
| **Substance Use** |  |
| **Alcohol Use, n (%)** |  |
| No use | 124 (56.6%) |
| Less than weekly | 38 (17.4%) |
| More than weekly | 25 (11.4%) |
| Daily or more | 12 (5.5%) |
| Not reported | 20 (9.1%) |
| **Benzodiazepine Use, Illicit, n (%)** |  |
| No use | 173 (79.0%) |
| Less than weekly | 1 (0.5%) |
| More than weekly | 0 (0%) |
| Daily or more | 0 (0%) |
| Not reported | 45 (20.5%) |
| **Benzodiazepine Use, Prescription n (%)** |  |
| No use | 157 (71.7%) |
| Less than weekly | 1 (0.5%) |
| More than weekly | 4 (1.8%) |
| Daily or more | 57 (26.0%) |
| Not reported | 0 (0%) |
| Cannabis Use, n (%) |  |
| No use | 153 (69.9%) |
| Less than weekly | 9 (4.1%) |
| More than weekly | 8 (3.7%) |
| Daily or more | 12 (5.5%) |
| Not reported | 37 (16.9%) |
| Nicotine Use, n (%) |  |
| No use | 160 (73.1%) |
| Less than weekly | 1 (0.5%) |
| More than weekly | 0 (0%) |
| Daily or more | 11 (5.0%) |
| Not reported | 47 (21.5%) |
| Inhalant Use, n (%) |  |
| No use | 151 (68.9%) |
| Less than weekly | 0 (0%) |
| More than weekly | 0 (0%) |
| Daily or more | 0 (0%) |
| Not reported | 68 (31.1%) |
| Opioid Use, Illicit, n (%) |  |
| No use | 179 (81.7%) |
| Less than weekly | 0 (0%) |
| More than weekly | 0 (0%) |
| Daily or more | 1 (0.5%) |
| Not reported | 39 (17.8%) |
| Opioid Use, Prescription, n (%) |  |
| No use | 206 (94.1%) |
| Less than weekly | 1 (0.5%) |
| More than weekly | 0 (0%) |
| Daily or more | 12 (5.5%) |
| Not reported | 0 (0%) |
| Other Substance Use, n (%) |  |
| No use | 148 (67.6%) |
| Less than weekly | 1 (0.5%) |
| More than weekly | 0 (0%) |
| Daily or more | 2 (0.9%) |
| Not reported | 68 (31.1%) |
| Psychedelic Use, n (%) |  |
| No use | 151 (68.9%) |
| Less than weekly | 1 (0.5%) |
| More than weekly | 0 (0%) |
| Daily or more | 0 (0%) |
| Not reported | 67 (30.6%) |
| Stimulant Use, Illicit, n (%) |  |
| No use | 174 (79.5%) |
| Less than weekly | 0 (0%) |
| More than weekly | 0 (0%) |
| Daily or more | 0 (0%) |
| Not reported | 45 (20.5%) |
| Stimulant Use, Prescription, n (%) |  |
| No use | 173 (79.0%) |
| Less than weekly | 0 (0%) |
| More than weekly | 0 (0%) |
| Daily or more | 46 (21.0%) |
| Not reported | 0 (0%) |

We performed ANCOVAs for each substance category to model the main effect of substance use frequency on change in PHQ-9 score controlling for age, sex, number of TMS sessions, and pre-TMS PHQ-9 score. For illicit benzodiazepines, illicit opioids, illicit stimulants, inhalants, other substance use, and psychedelics, fewer than four individuals reported any use. Therefore, it was determined there were insufficient data to support regression analyses for these substances. Across all other substance use categories, substance use frequency was not significantly associated with change in PHQ-9 score (all *p’s* > 0.05, Table 2). The Cohen’s d effect sizes ranged from 0.00 to 0.30, which would indicate a small effect of substance use on TMS response.

**Table 2.** ANCOVA Table of Substance Use and Antidepressant Response to TMS.

| Term | F-statistic | df | p-value | Cohen's d |
| --- | --- | --- | --- | --- |
| **Alcohol Use** |  |  |  |  |
| Pre-TMS PHQ-9 | 11.93 | 1 | 0.001 | 0.533 |
| Substance use frequency | 1.28 | 3 | 0.282 | 0.302 |
| Sex | 0.14 | 1 | 0.710 | 0.058 |
| Age | 2.25 | 1 | 0.135 | 0.231 |
| Total TMS sessions | 3.59 | 1 | 0.060 | 0.292 |
| **Benzodiazepine Use, Prescription** |  |  |  |  |
| Pre-TMS PHQ-9 | 15.48 | 1 | <0.001 | 0.575 |
| Substance use frequency | 1.36 | 3 | 0.255 | 0.295 |
| Sex | 0.24 | 1 | 0.628 | 0.072 |
| Age | 1.06 | 1 | 0.304 | 0.151 |
| Total TMS sessions | 8.91 | 1 | 0.003 | 0.437 |
| **Cannabis Use** |  |  |  |  |
| Pre-TMS PHQ-9 | 8.81 | 1 | 0.003 | 0.480 |
| Substance use frequency | 0.28 | 3 | 0.838 | 0.148 |
| Sex | 1.94 | 1 | 0.166 | 0.225 |
| Age | 1.63 | 1 | 0.204 | 0.206 |
| Total TMS sessions | 4.33 | 1 | 0.039 | 0.336 |
| **Nicotine Use** |  |  |  |  |
| Pre-TMS PHQ-9 | 6.79 | 1 | 0.010 | 0.433 |
| Substance use frequency | 1.05 | 2 | 0.352 | 0.241 |
| Sex | 1.53 | 1 | 0.218 | 0.205 |
| Age | 2.65 | 1 | 0.106 | 0.270 |
| Total TMS sessions | 5.33 | 1 | 0.022 | 0.383 |
| **Opioid Use, Prescription** |  |  |  |  |
| Pre-TMS PHQ-9 | 16.96 | 1 | <0.001 | 0.601 |
| Substance use frequency | 0.98 | 2 | 0.376 | 0.204 |
| Sex | 0.34 | 1 | 0.560 | 0.085 |
| Age | 0.82 | 1 | 0.368 | 0.132 |
| Total TMS sessions | 9.01 | 1 | 0.003 | 0.438 |
| **Stimulant Use, Prescription** |  |  |  |  |
| Pre-TMS PHQ-9 | 16.99 | 1 | <0.001 | 0.600 |
| Substance use frequency | 0.00 | 1 | 0.991 | 0.000 |
| Sex | 0.20 | 1 | 0.652 | 0.065 |
| Age | 1.35 | 1 | 0.247 | 0.169 |
| Total TMS sessions | 8.77 | 1 | 0.003 | 0.431 |

To compare individual levels of substance use frequency (i.e., daily use vs. no use), we modeled the effect of substance use frequency on change in PHQ-9 score controlling for age, sex, number of TMS sessions, and pre-TMS PHQ-9 score for each substance type. Across all substance use categories, substance use frequency was not associated with change in PHQ-9 score (all *p’s* > 0.05, Table 3). With the exception of less than weekly prescription benzodiazepine use (Est = −5.53, 95% CI [-18.23 to 7.17], p=0.39) and less than weekly prescription opioid use (Est = −6.98, 95% CI [-19.70 to 5.73], p=0.28), the range of plausible effects of substance use frequency on PHQ-9 change were between −5 and 5, suggesting substance use frequency was unlikely to have a meaningful clinical effect on antidepressant response to TMS. Pre-TMS PHQ-9 score was consistently associated with change in PHQ-9 score for each substance category, as patients with higher scores pre-TMS tended to have greater improvement after TMS. The results for each substance are detailed below:

**Table 3.** Models of Substance Use Levels and Antidepressant Response to TMS.

| Variable | Estimate | 95% Lower CI | 95% Upper CI | t | df | p-value | Cohen's d |
| --- | --- | --- | --- | --- | --- | --- | --- |
| **Alcohol Use** |  |  |  |  |  |  |  |
| Intercept | 5.07 | -2.08 | 12.22 | 1.40 | 168 | 0.16 | 0.216 |
| Pre-TMS PHQ-9 | -0.39 | -0.61 | -0.17 | -3.45 | 168 | <0.001 | -0.532 |
| Less than Weekly | -2.37 | -4.79 | 0.053 | -1.93 | 168 | 0.055 | -0.298 |
| More than Weekly | -0.85 | -3.78 | 2.07 | -0.58 | 168 | 0.57 | -0.089 |
| Daily or More | -0.019 | -3.88 | 3.85 | -0.0098 | 168 | 0.99 | -0.002 |
| Sex | 0.37 | -1.58 | 2.31 | 0.37 | 168 | 0.71 | 0.057 |
| Age | -0.049 | -0.11 | 0.015 | -1.51 | 168 | 0.14 | -0.233 |
| Total TMS Sessions | -0.12 | -0.25 | 0.0052 | -1.89 | 168 | 0.060 | -0.292 |
| **Benzodiazepine Use, Prescription** |  |  |  |  |  |  |  |
| Intercept | 6.16 | -0.32 | 12.63 | 1.88 | 187 | 0.062 | 0.275 |
| Pre-TMS PHQ-9 | -0.42 | -0.63 | -0.21 | -3.93 | 187 | <0.001 | -0.575 |
| Less than Weekly | -5.53 | -18.23 | 7.17 | -0.86 | 187 | 0.39 | -0.126 |
| More than Weekly | -6.48 | -13.85 | 0.90 | -1.73 | 187 | 0.085 | -0.253 |
| Daily or More | -0.82 | -2.96 | 1.31 | -0.76 | 187 | 0.45 | -0.111 |
| Sex | 0.45 | -1.38 | 2.29 | 0.49 | 187 | 0.63 | 0.072 |
| Age | -0.032 | -0.092 | 0.029 | -1.03 | 187 | 0.30 | -0.151 |
| Total TMS Sessions | -0.18 | -0.29 | -0.060 | -2.98 | 187 | 0.0032 | -0.436 |
| **Cannabis Use** |  |  |  |  |  |  |  |
| Intercept | 3.78 | -3.91 | 11.47 | 0.97 | 153 | 0.33 | 0.157 |
| Pre-TMS PHQ-9 | -0.35 | -0.59 | -0.12 | -2.97 | 153 | 0.0035 | -0.480 |
| Less than Weekly | 0.23 | -4.13 | 4.59 | 0.11 | 153 | 0.92 | 0.018 |
| More than Weekly | -1.79 | -6.47 | 2.89 | -0.76 | 153 | 0.45 | -0.123 |
| Daily or More | -1.07 | -4.88 | 2.75 | -0.55 | 153 | 0.58 | -0.089 |
| Sex | 1.44 | -0.60 | 3.48 | 1.39 | 153 | 0.17 | 0.225 |
| Age | -0.043 | -0.11 | 0.024 | -1.28 | 153 | 0.20 | -0.207 |
| Total TMS Sessions | -0.14 | -0.26 | -0.0069 | -2.08 | 153 | 0.039 | -0.336 |
| **Nicotine Use** |  |  |  |  |  |  |  |
| Intercept | 4.44 | -3.32 | 12.20 | 1.13 | 145 | 0.26 | 0.188 |
| Pre-TMS PHQ-9 | -0.31 | -0.55 | -0.076 | -2.61 | 145 | 0.010 | -0.433 |
| Less than Weekly | 4.39 | -8.38 | 17.17 | 0.68 | 145 | 0.50 | 0.113 |
| More than Weekly | - | - | - | - | - | - | - |
| Daily or More | -2.71 | -6.88 | 1.47 | -1.28 | 145 | 0.20 | -0.213 |
| Sex | 1.31 | -0.78 | 3.40 | 1.24 | 145 | 0.22 | 0.206 |
| Age | -0.057 | -0.13 | 0.012 | -1.63 | 145 | 0.11 | -0.271 |
| Total TMS Sessions | -0.16 | -0.29 | -0.023 | -2.31 | 145 | 0.022 | -0.384 |
| **Opioid Use, Prescription** |  |  |  |  |  |  |  |
| Intercept | 6.05 | -0.42 | 12.53 | 1.84 | 188 | 0.067 | 0.268 |
| Pre-TMS PHQ-9 | -0.43 | -0.64 | -0.23 | -4.12 | 188 | <0.001 | -0.601 |
| Less than Weekly | -6.98 | -19.70 | 5.73 | -1.08 | 188 | 0.28 | -0.158 |
| More than Weekly | - | - | - | - | - | - | - |
| Daily or More | -1.88 | -5.90 | 2.13 | -0.93 | 188 | 0.36 | -0.136 |
| Sex | 0.55 | -1.30 | 2.39 | 0.58 | 188 | 0.56 | 0.085 |
| Age | -0.028 | -0.089 | 0.033 | -0.90 | 188 | 0.37 | -0.131 |
| Total TMS Sessions | -0.18 | -0.29 | -0.061 | -3.00 | 188 | 0.003 | -0.438 |
| **Stimulant Use, Prescription** |  |  |  |  |  |  |  |
| Intercept | 6.28 | -0.20 | 12.76 | 1.91 | 189 | 0.057 | 0.278 |
| Pre-TMS PHQ-9 | -0.44 | -0.64 | -0.23 | -4.12 | 189 | <0.001 | -0.599 |
| Less than Weekly | - | - | - | - | - | - | - |
| More than Weekly | - | - | - | - | - | - | - |
| Daily or More | 0.012 | -2.17 | 2.19 | 0.011 | 189 | 0.99 | 0.002 |
| Sex | 0.42 | -1.42 | 2.26 | 0.45 | 189 | 0.65 | 0.065 |
| Age | -0.035 | -0.095 | 0.025 | -1.16 | 189 | 0.25 | -0.169 |
| Total TMS Sessions | -0.18 | -0.29 | -0.059 | -2.96 | 189 | 0.0035 | -0.431 |

PHQ-9: Patient Health Questionnaire-9; TMS: Transcranial Magnetic Stimulation. Substance use frequency is reported compared to “no use.” Dashes indicate empty categories.

### Alcohol

In the ANCOVA predicting change in depression symptom severity (PHQ-9) based on alcohol use frequency, pre-TMS PHQ-9, age, sex, and number of TMS sessions, only pre-TMS PHQ-9 (F=11.93, df=1, p=0.001) was a significant predictor (Table 2). In the multiple regression model predicting change in depression symptom severity (PHQ-9) based on level of alcohol use frequency (compared to no use), pre-TMS PHQ-9, age, sex, and number of TMS sessions, only pre-TMS PHQ-9 (Estimate = −0.39, t(168)=-3.45, p<.001) was a significant predictor (Figure 1, Table 3).

**Figure 1.**
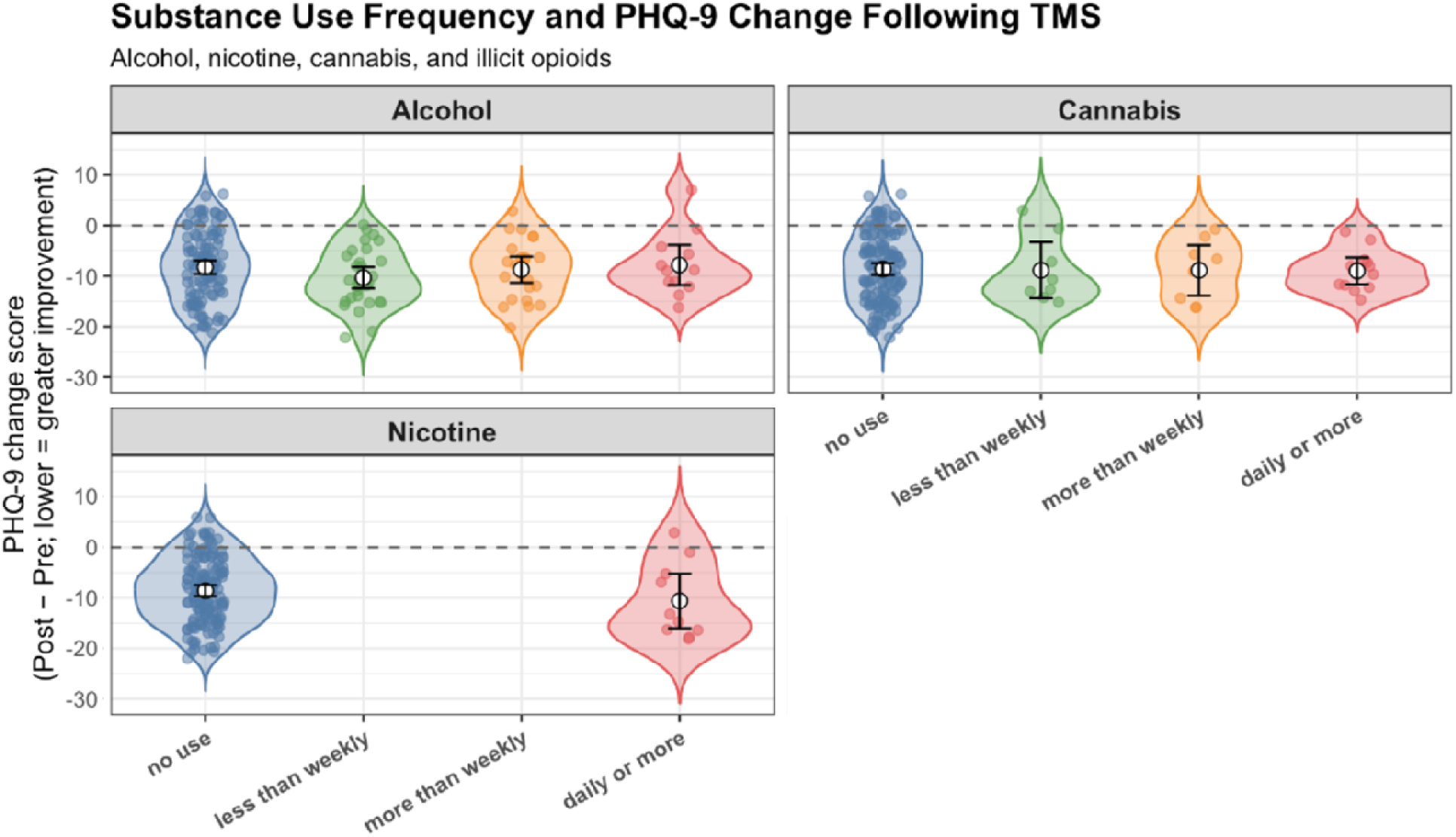
Associations Between Alcohol, Cannabis, and Nicotine Use and Response to TMS. We modeled change in depression symptom severity (PHQ-9) based on frequency of substance use, pre-TMS PHQ-9, age, sex, and number of TMS sessions. For each model based on alcohol, cannabis, and nicotine, substance use frequency did not predict antidepressant response to TMS (i.e., change in PHQ-9 score, ps>.05).

### Benzodiazepines

In the ANCOVA predicting change in depression symptom severity (PHQ-9) based on prescription benzodiazepine use frequency, pre-TMS PHQ-9, age, sex, and number of TMS sessions, only pre-TMS PHQ-9 (F=15.48, df=1, p<.001) was a significant predictor (Table 2). In the multiple regression model predicting depression symptom severity change based on level of prescription benzodiazepine use frequency (compared to no use), pre-TMS PHQ-9, age, sex, and number of TMS sessions, pre-TMS PHQ-9 (Estimate = −0.42, t(187)=-3.93, df=187.00, p<.001) and number of TMS sessions (Estimate = −0.18, t(187)= −2.98, p=0.0032) were significant predictors (Figure 2, Table 3).

**Figure 2.**
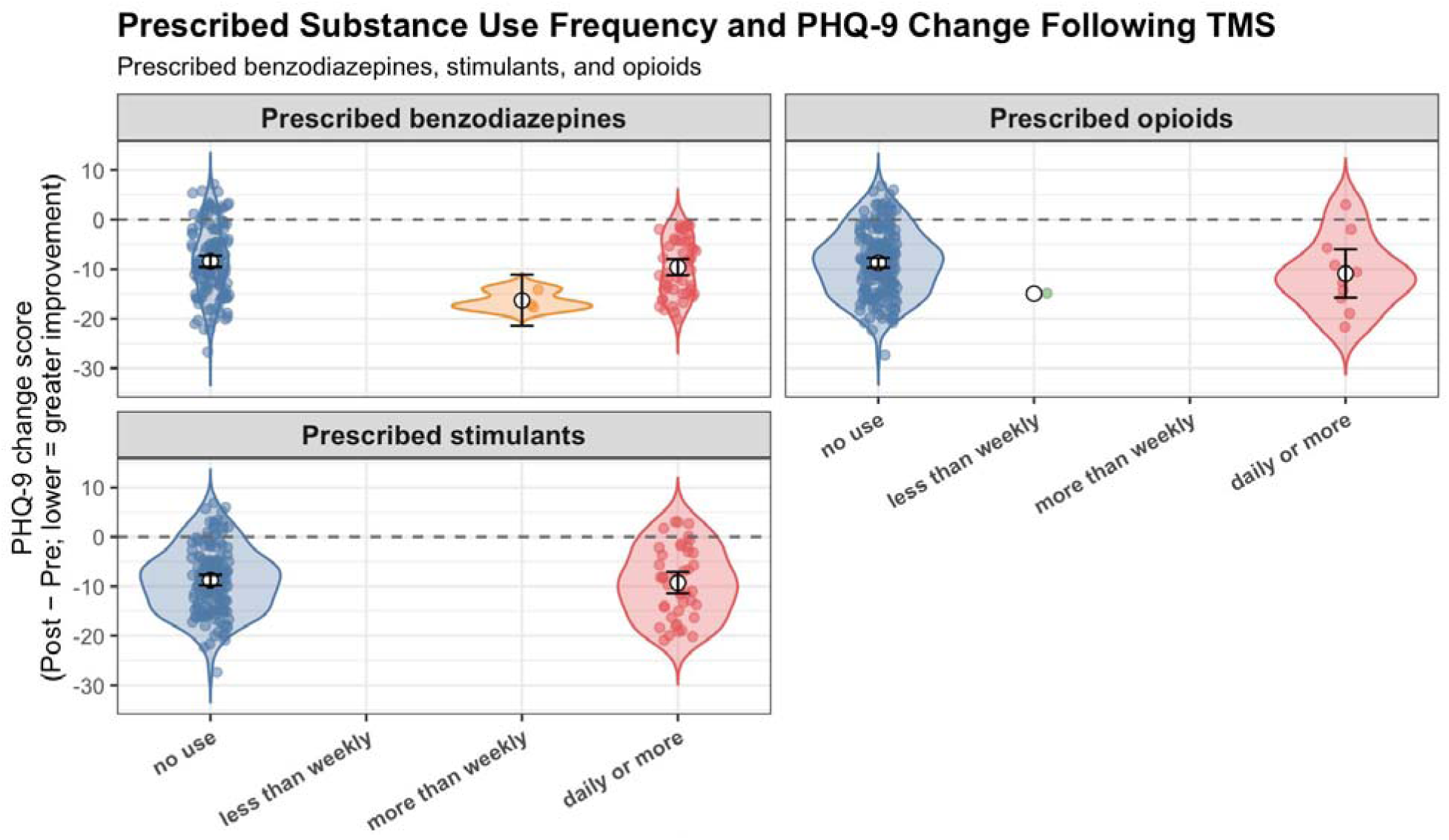
Associations Between Prescription Benzodiazepines, Opioids, and Stimulants and Response to TMS. We modeled change in depression symptom severity (PHQ-9) based on frequency of prescription substance use, pre-TMS PHQ-9, age, sex, and number of TMS sessions. For each model based on prescription benzodiazepine, opioid, and stimulant use, prescription substance use frequency did not predict antidepressant response to TMS (i.e., change in PHQ-9 score, ps>.05).

### Cannabis

In the ANCOVA predicting change in depression symptom severity (PHQ-9) based on cannabis use frequency, pre-TMS PHQ-9, age, sex, and number of TMS sessions, only pre-TMS PHQ-9 (F=8.81, df=1, p=0.003) was a significant predictor (Table 2). When we modeled depression symptom change based on level of cannabis use (compared to no use), pre-TMS PHQ-9, age, sex, and number of TMS sessions, pre-TMS PHQ-9 (Estimate = −0.35, t(153)=-2.97, p=0.0035) and number of TMS sessions (Estimate = −0.14, t(153)= −2.08, p=0.039) were significant predictors (Figure 1, Table 3).

### Nicotine

In the ANCOVA predicting change in depression symptom severity (PHQ-9) based on nicotine use frequency, pre-TMS PHQ-9, age, sex, and number of TMS sessions, only pre-TMS PHQ-9 (F=6.79, df=1, p=0.01) was a significant predictor (Table 2). In the multiple regression model predicting change in depression symptom severity based on level of nicotine use frequency (compared to no use), pre-TMS PHQ-9, age, sex, and number of TMS sessions, pre-TMS PHQ-9 (Estimate = −0.31, t(145)= −2.61, p=0.010) and number of TMS sessions (Estimate = −0.16, t(145)= −2.31, p=0.022) were significant predictors (Figure 1, Table 3).

### Opioids

In the ANCOVA predicting change in depression symptom severity (PHQ-9) based on prescription opioid use frequency, pre-TMS PHQ-9, age, sex, and number of TMS sessions, only pre-TMS PHQ-9 (F=16.96, df=1, p<0.001) was a significant predictor (Table 2). When we used multiple regression to model depression symptom change based on frequency of prescription opioid use, pre-TMS PHQ-9, age, sex, and number of TMS sessions, pre-TMS PHQ-9 (Estimate = −0.43, t(188)= −4.12, p<0.001) and number of TMS sessions (Estimate = −0.18, t(188)= −3.00, p=0.003) were significant predictors (Figure 2, Table 3).

### Stimulants

In the ANCOVA predicting change in depression symptom severity (PHQ-9) based on prescription stimulant use frequency, pre-TMS PHQ-9, age, sex, and number of TMS sessions, only pre-TMS PHQ-9 (F=16.99, df=1, p<.001) was a significant predictor (Table 2). When we used multiple regression to model change in depression symptom severity based on level of prescription stimulant use (compared to no use), pre-TMS PHQ-9, age, sex, and number of TMS sessions, pre-TMS PHQ-9 (Estimate = −0.44, t(189)= −4.12, p<.001) and number of TMS sessions (Estimate = −0.18, t(189)= −2.96, p=0.0035) were significant predictors (Figure 2, Table 3).

## Discussion

In this retrospective cohort study, we assessed the impact of concurrent substance use on change in PHQ-9 score to determine whether substance impacts the response to TMS for the treatment of depression. We found that substance use, specifically alcohol, cannabis, nicotine, prescribed stimulant, prescribed benzodiazepine, and opioid use (prescribed and illicit) was not significantly associated with response to TMS, and the effect size estimates were below the threshold of clinical significance. Given the high rates of co-occurring substance use and MDD, these results help to clarify that concurrent substance use does not impact the efficacy of TMS for treating symptoms of depression and may be useful in informing clinical decision-making.

Concurrent alcohol use, the most commonly reported substance in our sample, was not associated with reduced efficacy of TMS in alleviating depressive symptoms in the current sample. Alcohol use is common among individuals diagnosed with MDD, where alcohol consumption is used as a means to cope with symptoms of depression (Holahan, Moos, Holahan, Cronkite, & Randall, 2001; Kenney, Jones, & Barnett, 2015). Indeed, consistent with the self-medication hypothesis where drinking is motivated by a desire to reduce negative affect (Magee & Connell, 2021), depressive symptoms have been positively related to weekly alcohol consumption (Villarosa, Messer, Madson, & Zeigler-Hill, 2018). Accordingly, over 30% of the current sample reported drinking alcohol, although only 11% of the sample reported drinking more than weekly. Thus, while MDD frequently co-occurs with alcohol use disorder (AUD; Brière, Rohde, Seeley, Klein, & Lewinsohn, 2014; McHugh & Weiss, 2019), most participants in the present study would likely not have met diagnostic criteria for AUD based on their reported frequency of use. This distinction is clinically important because many of the factors that may complicate TMS treatment, such as alcohol withdrawal syndrome (Rossi et al., 2021), are more commonly associated with problematic alcohol use than with moderate drinking. As such, consideration of alcohol use in the context of TMS treatment should extend beyond simple presence or absence of consumption and include the frequency and severity of use. Nevertheless, our findings suggest that alcohol consumption, including weekly or daily use, may not significantly influence response to TMS treatment for depression.

Concurrent prescribed benzodiazepine use was also not associated with reduced efficacy of TMS for depression. The high prevalence of benzodiazepine use in individuals with MDD is unsurprising, as these medications are frequently prescribed for the treatment of anxiety, which co-occurs with MDD in more than 50% of cases (Davies et al., 2023; Kessler et al., 2012). Given the frequency with which prescribed benzodiazepines are used by this sample, it is encouraging that their use did not appear to diminish the antidepressant efficacy of TMS. Although early TMS studies often required participants to undergo medication washout prior to treatment, more recent studies have generally included patients who remain stable on their medications throughout treatment (Chen et al., 2025). Indeed, while previous work has suggested that GABA-ergic signalling crucial to the therapeutic effects of TMS would be mitigated by benzodiazepine use (Hunter et al., 2019), findings regarding the impact of concurrent benzodiazepine use on TMS outcomes have been mixed, with the overall literature increasingly supportive of their concurrent use. For example, analyzes of pooled data by Fitzgerald, Daskalakis, and Hoy (2020) found no significant differences in changes in depression rating scores between patients taking benzodiazepines and those who were not. Similarly, a recent review by Tran et al. (2024) found no evidence that benzodiazepine use negatively affected TMS response in depression. Moreover, while Hunter et al. (2019) observed reduced response rates among participants taking benzodiazepine, the findings did not remain significant after correction for multiple comparisons, and the authors emphasized the need for replication. Likewise, Deppe et al. (2021) reported that lorazepam use was associated with a reduced response to TMS, although all patients experienced improvements in depressive symptoms over the course of treatment. Thus, consistent with existing reviews, our findings suggest that benzodiazepine use, even at daily levels, does not significantly attenuate the antidepressant effects of TMS in a naturalistic clinical sample and support the continued inclusion of patients receiving benzodiazepines in routine clinical TMS practice.

Other classes of substances that were more commonly reported were also not significantly associated with PHQ-9 scores, including prescribed stimulants and cannabis. Psychostimulants are commonly used as adjunctive treatments for MDD (McIntyre et al., 2017), with evidence suggesting that their efficacy may be greater when combined with traditional antidepressant medications (Sadowska et al., 2026). In the present study, the use of prescribed stimulants during TMS treatment did not negatively affect antidepressant outcomes, findings that are consistent with two recent studies that found no detrimental effect of psychostimulant use on TMS outcomes for depression (Carr, Barbour, Harris, & Camprodon, 2025; Wilke et al., 2022). Aligning with other substances, cannabis use also did not attenuate response to TMS in our sample, directly supporting the work of Shenasa et al. (2023) and preliminary work by Tirrell et al. (2026) that found no effect of cannabis use on PHQ-9 outcomes.

The present findings thus contribute to a growing body of evidence suggesting that concurrent low to moderate substance use during TMS treatment for depression may not significantly impact treatment outcomes. Work by Gonzalez et al. (2026), conducted in a geographically distinct population with comparable low to moderate levels of alcohol, cannabis, and nicotine use, found that this recreational pattern of substance use was not associated with antidepressant response to TMS, consistent with our own findings. However, a limitation of this previous work was that it included limited information on frequency of substance use. Here, we extend this literature with more granular data on substance use frequency and demonstrate that regular prescription medication use, and infrequent use of other illicit substances also do not appear to attenuate the antidepressant effects of TMS. Collectively, these findings suggest that recreational use of most substances is unlikely to meaningfully affect improvements in depressive symptoms. It is important to note that participants in the present study were not engaging in regular or heavy substance use. This pattern may reflect existing clinical practices in which individuals with more frequent substance use are either less likely to be referred for TMS or are encouraged to reduce their use before treatment initiation, although this interpretation remains speculative. Nevertheless, the accumulating evidence from studies of patients with low to moderate substance use may help inform clinical decision-making regarding the appropriateness of TMS for individuals who engage in occasional substance use.

Our findings have important clinical implications. Existing clinical guidelines have recommended abstinence from substances prior to a course of TMS for MDD (Tang et al., 2025). Our work and others’ (Carr et al., 2025); Gonzalez et al. (2026); Shenasa et al. (2023); Tirrell et al. (2026); (Wilke et al., 2022) suggests that low to moderate substance use does not impact antidepressant response to TMS. TMS is a highly effective treatment for depression yet remains inaccessible to many individuals due to insurance coverage or geographic accessibility. Therefore, low level substance use should not pose yet another barrier to treatment for a population that so desperately needs it.

However, our analysis has several limitations. Most importantly, because our sample had low to moderate substance use, we cannot comment on the efficacy of TMS for depression in individuals with a co-occurring substance use disorder. However, TMS is FDA-cleared for smoking cessation (Zangen et al., 2021) and has accumulating evidence supporting its use for substance use disorders (Mehta et al., 2023). Another limitation is that data on substance use frequency was collected from a standard TMS consultation by a psychiatrist, rather than using detailed instruments. Therefore, we lack more granular information on level of substance dependence, use patterns, craving, and withdrawal as well as biochemical verification of substance use (e.g., urine toxicology). Future work should examine more detailed substance use information in larger, more diverse samples.

## Conclusion

Our retrospective cohort study analyzing how substance use impacts change depression symptoms pre-and post-TMS showed that frequency of substance use status did not influence response to TMS. This knowledge may help guide providers and patients alike in making treatment decisions and expanding access to this highly effective treatment.

## Data Availability

Data may be available upon reasonable request.

## Acknowledgements

This work was funded by National Institutes of Health K23DA059690 to Dr. Ward.

